# Lysosomal degradation of PD-L1 is associated with immune-related adverse events during anti-PD-L1 immunotherapy in NSCLC patients

**DOI:** 10.1101/2024.01.21.24301536

**Authors:** Takeru Kashiwada, Ryotaro Takano, Fumihiko Ando, Shoko Kuroda, Yoshishige Miyabe, Tomoko Asatsuma-Okumura, Masaaki Hashiguchi, Yoshikazu Kanazawa, Hiroshi Yoshida, Masahiro Seike, Akihiko Gemma, Yoshiko Iwai

**Affiliations:** Department of Pulmonary Medicine and Oncology, Graduate School of Medicine, Nippon Medical School, Tokyo, Japan; Department of Cell Biology, Institute for Advanced Medical Sciences, Graduate School of Medicine, Nippon Medical School, Tokyo, Japan; Department of Gastrointestinal and Hepato-Biliary-Pancreatic Surgery, Graduate School of Medicine, Nippon Medical School, Tokyo, Japan; Department of Immunology and Parasitology, St. Marianna University School of Medicine, Kanagawa, Japan

**Keywords:** Soluble PD-L1, inflammation, macrophage, lysosomal degradation, immune-related adverse event

## Abstract

**BACKGROUND:** Immune checkpoint inhibitors (ICIs) can induce immune-related adverse events (irAEs). However, the biological mechanisms underlying irAEs are not well understood. We previously developed an ELISA system to specifically detect soluble PD-L1 (sPD-L1) with PD-1-binding capacity (bsPD-L1). Here, we investigated the relationship between sPD-L1 and bsPD-L1 in gastric cancer (GC) and non-small cell lung cancer (NSCLC) patients treated with PD-1/PD-L1 blockade and their association with irAEs.

**METHODS:** We examined sPD-L1, bsPD-L1, matrix metalloproteinases (MMPs), and proinflammatory cytokine levels by ELISA in plasma samples from 117 GC patients prior to surgery and 72 NSCLC patients prior to and at 2 months after ICI treatment (anti-PD-1, n = 48; anti-PD-L1, n = 24). In mice treated with anti-PD-1/PD-L1 antibodies (Abs), sPD-L1 levels and localization of Abs were examined by ELISA and immunohistochemistry, respectively.

**RESULTS:** sPD-L1 was detected with higher frequency in GC patients (76.9%) than in NSCLC patients (8.3%), whereas bsPD-L1 was detected with similar frequencies in GC and NSCLC patients (14.5% and 22.2%, respectively). sPD-L1 levels were correlated with IL-1α, IL-1β, TNF-α, and IL-6 levels, while bsPD-L1 levels were correlated with MMP13, MMP3, and IFN-γ levels. In NSCLC patients, anti-PD-L1, but not anti-PD-1, treatment increased sPD-L1, which was associated with irAE development, but not with clinical outcomes. In mice, trafficking of anti-PD-L1 Abs to lysosomes in F4/80^+^ macrophages resulted in sPD-L1 production, which was suppressed by treatment with lysosomal degradation inhibitor chloroquine and macrophage depletion.

**CONCLUSIONS:** Anti-PD-L1-mediated lysosomal degradation induces sPD-L1 production, which can serve as an indicator to predict irAE development during anti-PD-L1 treatment.

## Background

Immune checkpoint inhibitors (ICIs) targeting programmed cell death 1 (PD-1) and its ligand, PD-L1, can induce durable anti-tumor response, but can also cause immune-related adverse events (irAEs) ^1^ ^2^. irAEs affect almost any organs and are sometimes fatal. Additionally, the onset of irAEs is unpredictable. Therefore, biomarkers to predict irAE occurrence are urgently needed. Our early studies demonstrated the inhibitory role of PD-1/PD-L1 signaling in anti-tumor immune response^3–7^. PD-L1 is constitutively expressed on endothelial cells and macrophages in peripheral tissues^4^. Proinflammatory cytokines such as tumor necrosis factor (TNF)-α and interferon (IFN)-γ induce PD-L1 expression in various cell types, including immune, parenchymal, and tumor cells^4^. Upon ligation with PD-L1, PD-1 inhibits T cell proliferation and effector functions^3^ ^4^. Both anti-PD-1 and anti-PD-L1 monoclonal antibodies (mAbs) enhance the T cell response, but the difference in their mechanism of action is not fully understood.

In addition to the membrane-bounded form, PD-L1 also has a soluble form (sPD-L1)^8^ ^9^. Many studies have conflicting results on the function of sPD-L1^10–12^. sPD-L1 is produced via several mechanisms^9^ ^13–16^, including RNA splicing, exosomal secretion, and proteolytic cleavage by matrix metalloproteinases (MMPs). We previously developed an ELISA system to specifically detect PD-1-binding sPD-L1 (bsPD-L1)^17^, and found that bsPD-L1 can function as an endogenous PD-1 blocker (preprint in medRxiv/2023/300672). Plasma bsPD-L1 is strongly correlated with MMP13 levels. bsPD-L1 and MMP13 levels are associated with intra-tumoral T cell infiltration and loss of extracellular matrix integrity, respectively, in the tumor microenvironment. The levels of MMP13 and its activator, MMP3, change during ICI treatment. Furthermore, the combination of bsPD-L1 and MMPs may serve as a non-invasive tool to predict the efficacy of ICIs. However, the relationship between bsPD-L1 and sPD-L1, and their difference in clinical significance remain unknown.

In this study, we investigated plasma bsPD-L1 and sPD-L1 levels in GC and NSCLC patients treated with PD-1/PD-L1 blockade. We found that anti-PD-L1, but not anti-PD-1, treatment induced sPD-L1 production, which may serve as a non-invasive tool to predict irAEs.

## Methods

### Patients and specimens

This study included 117 patients diagnosed with GC between 2017 and 2020 at the Department of Gastrointestinal and Hepato-Biliary-Pancreatic Surgery in Nippon Medical School Hospital, Japan. Blood samples were collected from patients before surgery.

The study also included 72 patients diagnosed with NSCLC between 2017 and 2019 at the Department of Pulmonary Medicine and Oncology in Nippon Medical School Hospital, Japan. Blood was collected from patients prior to and at 2 months after the initiation of checkpoint immunotherapy (nivolumab, n = 20; pembrolizumab, n = 28; atezolizumab, n = 16; durvalumab, n=8). Nivolumab (3 mg/kg, every 2 weeks), pembrolizumab (200 mg/body, every 3 weeks), or atezolizumab (1200 mg/body, every 3 weeks) was administered until disease progression or unacceptable adverse events in a clinical setting. Durvalumab (10 mg/kg, every 2 weeks) was administered after curative chemoradiotherapy.

RECIST v1.1 was used to assess the efficacy of immunotherapy. Patients with progressive disease (PD) who were not evaluable for response by RECIST were determined by the treating physician as PD. We assessed disease control [DC; complete response (CR) + partial response (PR) + stable disease (SD)] at 2 months.

Baseline clinical and demographic data were collected from patient medical records. The study protocols (B-2019-005, and 28-09-646) were reviewed and approved by the Ethics Committee of Nippon Medical School. All participants provided written informed consent. This study was conducted in accordance with the Declaration of Helsinki.

### Animal experiments

C57BL/6 (B6) mice were purchased from Oriental Yeast Co., Ltd. (Tokyo, Japan). Mice were intravenously injected with 100 μg of anti-PD-1 (clone: J43, Invitrogen, Waltham, MA, USA, #16-9985-85) or anti-PD-L1 (clone: MIH5, Invitrogen #16-5982-85) mAbs, and then blood was collected at several timepoints. To inhibit lysosomal degradation, mice were intraperitoneally injected with chloroquine (100 mg/kg/day, Sigma-Aldrich, St. Louis, MO, USA, #C6628) for 3 days prior to Ab administration. To deplete macrophages, mice were intravenously injected with clodronate-containing or control liposomes (12.5 mg/kg, Hygieia Bioscience, Osaka, Japan, #16001003) at 24 hours prior to Ab administration. All mice were maintained at the animal facility of Nippon Medical School. All animal experiments were approved by the Animal Care and Use Committee of Nippon Medical School (2020-018).

### Enzyme-linked immunosorbent assay (ELISA)

Human sPD-L1 and bsPD-L1 concentrations were measured as described previously ^17^. Human MMP3, MMP9, MMP13, IL-1α, IL-1β, IL-6, TNF-α, and IFN-γ, and mouse sPD-L1 concentrations were determined using ELISA kits (R&D Systems, Minneapolis, MN, USA, #DY513, #DY911, #DY511, #DY200, #DY201, #DY206, #DY210, #DY285B, and #DY1019, respectively), in accordance with the manufacturer’s instructions.

### Immunohistochemistry

Mice were intravenously injected with 100 μg of Alexa 488-labeled mAb, and then organs were harvested 24 h later. Cryosections were fixed and permeabilized by Cytofix/Cytoperm solution (BD Biosciences, Franklin Lakes, NJ, USA, #554722) and stained with anti-B220-PE (BioLegend, San Diego, CA, USA, #103208) and anti-F4/80-Alexa Fluor 647 (BioLegend, #123122) mAbs. To examine subcellular localization of mAbs, cryosections were stained with anti-Rab7 (Abcam, Cambridge, UK, #ab137029) or anti-LAMP1 (Abcam #ab24170) Abs in combination with anti-F4/80-Alexa Fluor 647, followed by anti-rabbit IgG-Cy3 (Jackson ImmunoResearch, West Grove, PA, USA, #711-166-152). The sections were mounted with Prolong Gold with DAPI (Invitrogen, #P36931) and analyzed by confocal microscopy (Olympus, Tokyo, Japan, #FV1200). Images were processed using Imaris software, version 9.7.1(Oxford Instruments, Abingdon, UK).

### Statistical analysis

Mann-Whitney U test or Student’s t-test was used to compare numerical variables, and Pearson’s chi-squared test was used to compare categorical variables. Receiver operator characteristic (ROC) curve, along with area under the ROC (AUC), was used to assess the discriminatory ability of the numerical variables. All tests were two-tailed, and p-values < 0.05 were considered statistically significant. All statistical analyses were performed using JMP software, version 13 (SAS Institute, Cary, NC, USA) and Prism software, version 8 (GraphPad, San Diego, CA, USA).

## Results

### Detection of sPD-L1 with higher frequency in GC patients than in NSCLC patients

We examined baseline sPD-L1 and bsPD-L1 levels in plasma samples from 117 GC patients and 72 NSCLC patients (Figure 1A). sPD-L1 was detected with higher frequency in GC patients (90/117, 76.9%) than in NSCLC patients (6/72, 8.3%), whereas bsPD-L1 was detected with similar frequencies in GC patients (17/117, 14.5%) and NSCLC patients (17/72, 22.2%). The average sPD-L1 level in GC patients was higher than that in NSCLC patients (693 ± 2668 and 125 ± 837 pg/mL, respectively), while the average bsPD-L1 level in NSCLC patients was higher than that in GC patients (2237 ± 7960 and 216 ± 823 pg/mL, respectively). We found a stronger correlation between sPD-L1 and bsPD-L1 levels in GC patients than in NSCLC patients (correlation coefficient [r] = 0.8105 and r = 0.4232, respectively; Figure 1B). These results suggest different expression patterns of sPD-L1 and bsPD-L1 between GC and NSCLC.

**Figure 1.**
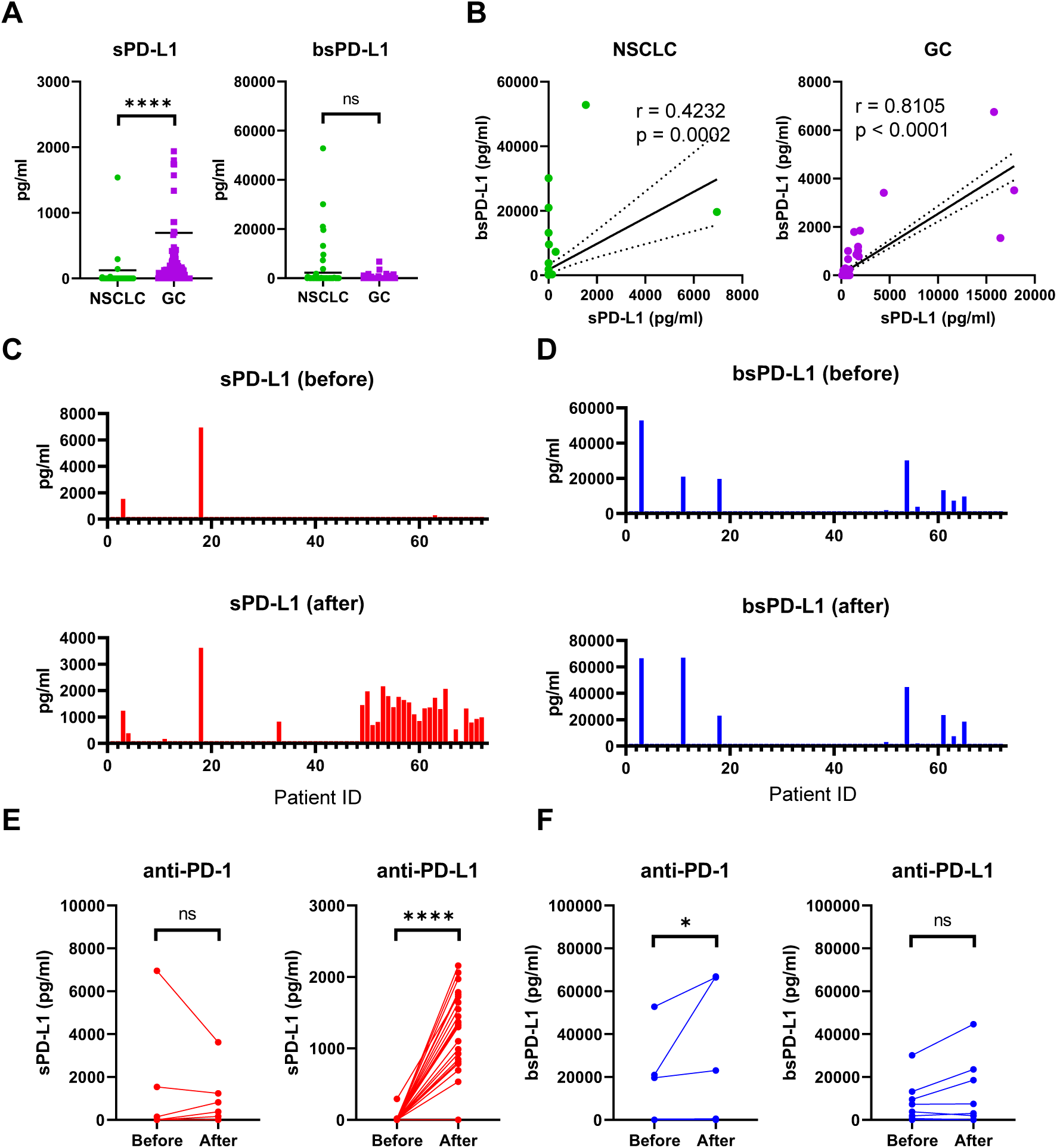
Detection of sPD-L1 and bsPD-L1 in the plasma of GC and NSCLC patients. (A) Baseline sPD-L1 and bsPD-L1 levels in plasma samples from GC patients (n = 117) and NSCLC patients (n = 72). (B) Correlation between baseline sPD-L1 and bsPD-L1 levels in GC and NSCLC patients. r indicates the correlation coefficient. (C) sPD-L1 levels and (D) bsPD-L1 levels in plasma samples from NSCLC patients prior to and at 2 months after ICI treatment. (E) Kinetic change of sPD-L1 and (F) bsPD-L1 levels in NSCLC patients prior to and at 2 months after anti-PD-1 (n = 48) or anti-PD-L1 (n = 24) treatment. r indicates the correlation coefficient. Horizontal lines indicate the mean. Statistical significance was calculated using the Mann– Whitney U test (A, E, and F). *p < 0.05; ****p < 0.0001; ns, not significant.

### Increased sPD-L1 levels during anti-PD-L1 treatment in NSCLC patients

Next, we investigated kinetic changes of sPD-L1 and bsPD-L1 in NSCLC patients during ICI treatment. Pretreatment patient characteristics are summarized in Supplementary Table 1. No significant differences were found in any of the variables between anti-PD-1- and anti-PD-L1-treated groups. At 2 months of treatment, sPD-L1^+^ patients were increased (Figure 1C), while bsPD-L1^+^ patients were unchanged (Figure 1D). Subgroup analysis revealed that anti-PD-L1 treatment increased sPD-L1 levels, but not bsPD-L1 levels (Figure 1E and 1F).

### Correlation between sPD-L1 and proinflammatory cytokine levels

It has been reported that PD-L1 is selectively cleaved by MMP13 and MMP9, in vitro^13^. We investigated the relationship between baseline MMPs and sPD-L1 or bsPD-L1 in NSCLC patients (Figure 2A). MMP13 levels were strongly corelated with bsPD-L1 levels, but weakly correlated with sPD-L1 levels. MMP3 levels were moderately correlated with bsPD-L1, but not with sPD-L1. MMP9 levels were not correlated with either sPD-L1 or bsPD-L1. These results suggest that MMP13 may be involved in the generation of bsPD-L1 rather than sPD-L1.

**Figure 2.**
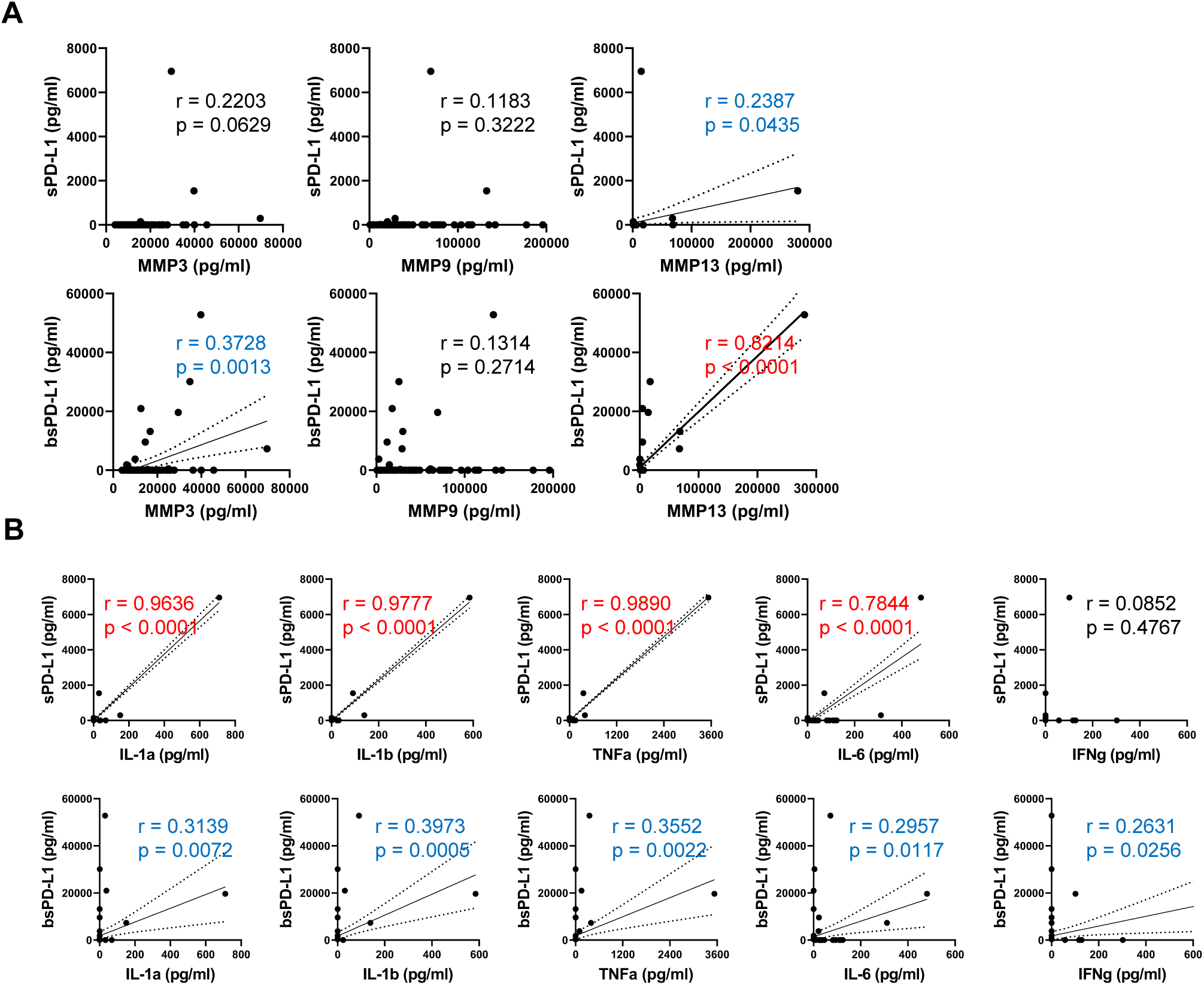
Correlation between baseline sPD-L1 and proinflammatory cytokine levels in NSCLC patients. (A) Correlation between baseline MMPs (MMP3, MMP9, and MMP13) and sPD-L1 or bsPD-L1 levels in NSCLC patients (n = 72). (B) Correlation between baseline proinflammatory cytokines (IL-1α, IL-1β, TNF-α, IL-6, and IFN-γ) and sPD-L1 or bsPD-L1 levels in NSCLC patients (n = 72). r indicates the correlation coefficient.

We investigated the relationship between proinflammatory cytokines and sPD-L1 or bsPD-L1 levels (Figure 2B). sPD-L1 levels were strongly correlated with IL-1α, IL-1β, TNF-α, and IL-6, whereas bsPD-L1 levels were weakly correlated with these cytokines, suggesting that inflammation may be involved in the generation of sPD-L1 rather than bsPD-L1. Furthermore, IFN-γ levels were weakly correlated with bsPD-L1 levels (r = 0.2631), but not with sPD-L1 levels, suggesting that bsPD-L1, but not sPD-L1, may regulate T cell response.

### Internalization of anti-PD-L1 mAb by mouse F4/80^+^ macrophages

To examine the mechanism of sPD-L1 production during immunotherapy, mice were treated with anti-PD-1 or anti-PD-L1 mAb (Figure 3A). Consistent with NSCLC patients, sPD-L1 levels were increased in mice treated with anti-PD-L1 mAb, but not with anti-PD-1 mAb. sPD-L1 levels began to increase at day 1, reached a peak at day 3 and remained detectable at day 7 of anti-PD-L1 mAb administration.

**Figure 3.**
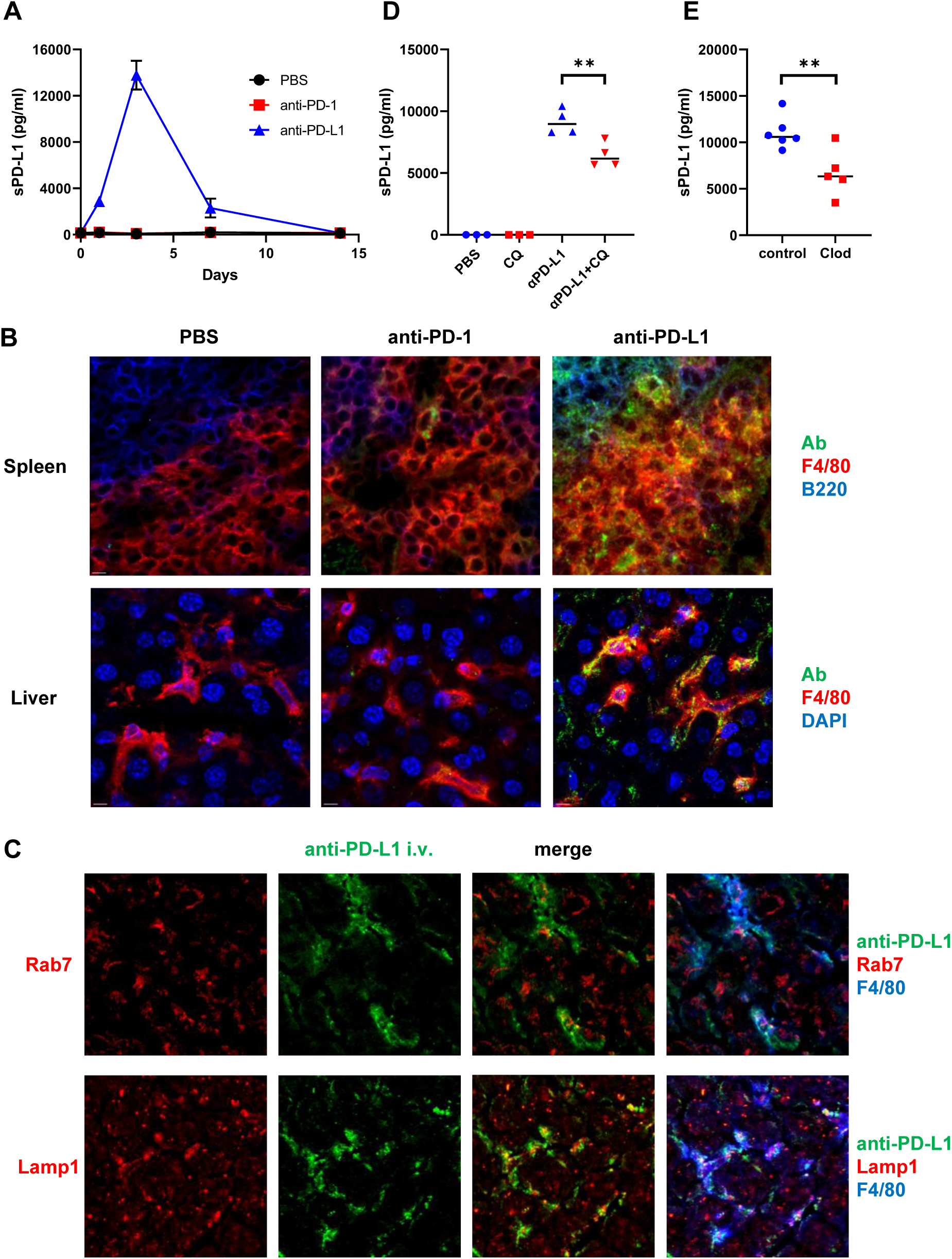
Intracellular trafficking of anti-PD-L1 mAb in mouse F4/80^+^ macrophages. (A) Mice were intravenously injected with PBS, anti-PD-1, or anti-PD-L1 mAb, and then blood was collected at the indicated time points. sPD-L1 levels were analyzed by ELISA. (B) Mice were intravenously injected with PBS, Alexa 488-labeled anti-PD-1, or anti-PD-L1 mAb (green), and then organs were harvested 24 h later. Spleen sections were stained with anti-F4/80 (red) and anti-B220 (blue) mAbs. Liver sections were stained with anti-F4/80 mAb (red) and DAPI (blue). (C) Mice were intravenously injected with Alexa 488-labeled anti-PD-L1 mAb (green), and then organs were harvested 24 h later. Liver sections were stained with anti-Rab7 or anti-LAMP1 Ab (red) in combination with anti-F4/80 mAb (blue). (D) Mice were treated with chloroquine (CQ) for 3 days and then intravenously injected with anti-PD-L1 mAb. Blood samples were collected 24 h later and sPD-L1 levels were analyzed by ELISA. (E) Mice were intravenously injected with clodronate-containing or control liposomes and then treated with anti-PD-L1 mAb. Blood samples were collected 24 h later, and sPD-L1 levels were analyzed by ELISA. Horizontal lines indicate the mean. Statistical significance was calculated using the Student’s t-test (D and E). **p < 0.01.

To investigate the tissue distribution of anti-PD-L1 mAb in vivo, mice were treated with fluorescently labeled anti-PD-L1 mAb. Splenic F4/80^+^ macrophages accumulated intracellular fluorescence (Figure 3B). The anti-PD-L1 mAb was localized predominantly inside the cells, with little remaining on the cell surface. However, the fluorescently labeled anti-PD-1 mAb remained almost exclusively on the surface of splenic F4/80^+^macrophages. The internalization of anti-PD-L1 mAb with punctate staining was also observed in liver and bone marrow F4/80^+^ macrophages (Figure 3B and Supplementary Figure 1). These results suggest that the injected anti-PD-L1 mAb was internalized by macrophages in various tissues.

### Macrophages as the source of sPD-L1 during anti-PD-L1 treatment

To investigate the subcellular localization of anti-PD-L1 mAb, mice were injected with the fluorescently labeled anti-PD-L1 mAb. After 24 hours, liver sections were stained with anti-Rab7 or anti-Lamp1 Ab (Figure 3C). Rab7 and Lamp1 are markers of late endosome and lysosome, respectively. Anti-PD-L1 staining was highly punctate in F4/80^+^macrophages. Some co-localization was observed between anti-PD-L1 and anti-Lamp1 Abs, suggesting that the anti-PD-L1 mAb may traffic to lysosomes in F4/80^+^ macrophages after internalization.

Ab binding to cell surface molecules often triggers endocytosis of antigen/Ab complexes, which are delivered to lysosomes for degradation^18^. To examine whether lysosomal degradation might be involved in sPD-L1 production, mice were treated with chloroquine prior to the anti-PD-L1 mAb administration. sPD-L1 levels were significantly decreased in chloroquine-treated mice compared with untreated mice (Figure 3D), suggesting that lysosomal degradation plays an important role in sPD-L1 production.

To examine whether macrophages might be involved in sPD-L1 production, mice were treated with clodronate-containing liposomes to delete macrophages prior to the anti-PD-L1 mAb administration. sPD-L1 levels were lower in mice treated with clodronate-containing liposomes than in those treated with control liposomes (Figure 3E), suggesting that macrophages are the source of sPD-L1 during anti-PD-L1 treatment.

### Elevated sPD-L1 in NSCLC patients with irAEs during anti-PD-L1 treatment

We investigated whether the sPD-L1 change was associated with clinical response to ICIs in NSCLC patients (Figure 4A and 4B). At 2 months of treatment, 33 of 48 anti-PD-1-treated patients and 17 of 24 anti-PD-L1-treated patients obtained DC. No significant difference was found in the sPD-L1 change between patients with DC and PD in both treatment groups. We also examined the association of the sPD-L1 change with irAE development during ICI treatment (Figure 4C and 4D). After ICI treatment, 20 of 48 anti-PD-1-treated patients and 11 of 24 anti-PD-L1-treated patients developed irAEs. Patients with irAEs experienced significantly greater increase in sPD-L1 than patients without irAEs during anti-PD-L1 treatment (p = 0.0321), but not during anti-PD-1 treatment.

**Figure 4.**
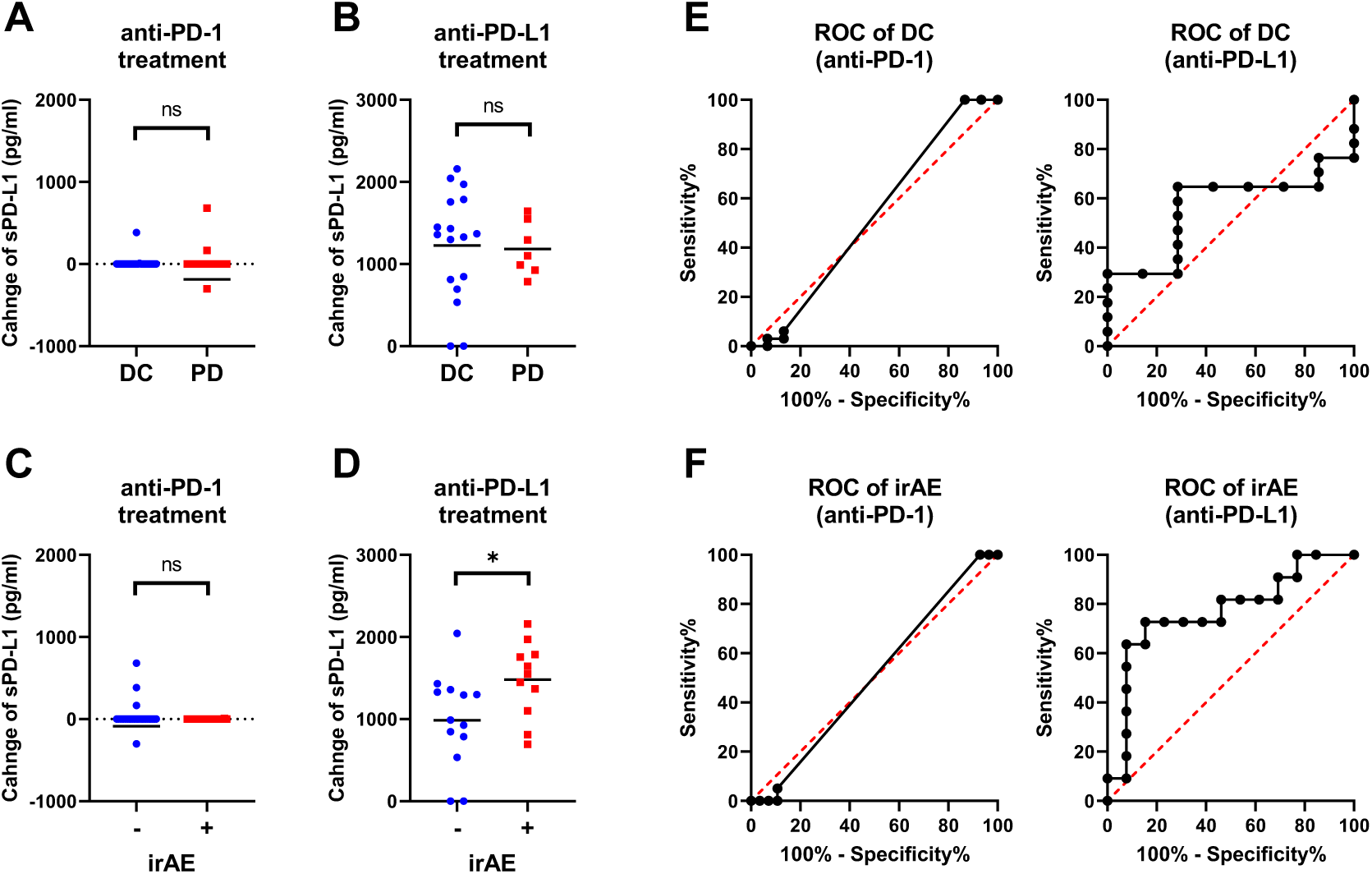
Increased sPD-L1 levels in NSCLC patients with irAEs during anti-PD-L1 treatment. (A) Comparison of the sPD-L1 change between patients with disease control (DC, n = 33) and progressive disease (PD, n = 15) at 2 months of anti-PD-1 treatment. (B) Comparison of the sPD-L1 change between patients with DC (n = 17) and PD (n = 7) at 2 months of anti-PD-L1 treatment. (C) Comparison of the sPD-L1 change between patients with irAEs (n = 20) and without irAEs (n = 28) during anti-PD-1 treatment. (D) Comparison of the sPD-L1 change between patients with irAEs (n = 11) and without irAEs (n = 13) during anti-PD-L1 treatment. (E) ROC curve analysis of the sPD-L1 change to predict DC in patients with anti-PD-1 or anti-PD-L1 treatment. (F) ROC curve analysis of the sPD-L1 change to predict irAEs in patients with anti-PD-1 or anti-PD-L1 treatment. Horizontal lines indicate the mean. Statistical significance was calculated using the Student’s t-test (A, B, C, and D). *p < 0.05; ns, not significant.

We performed ROC analysis to assess the discriminatory ability of the sPD-L1 change during anti-PD-L1 treatment. The AUC of the sPD-L1 change to predict DC was 0.563, indicating a low discriminatory ability of change in sPD-L1 for clinical outcomes (Figure 4E). Conversely, the AUC of the sPD-L1 change to predict irAE was 0.769, demonstrating a high discriminatory ability (Figure 4F). These results suggest that the potential utility of sPD-L1 change to predict irAEs during anti-PD-L1 treatment.

## Discussion

In this study, we investigated the relationship between sPD-L1 and bsPD-L1 in GC and NSCLC patients. sPD-L1 was detected with higher frequency in GC patients than in NSCLC patients, whereas bsPD-L1 was detected with similar frequencies in GC and NSCLC patients. In NSCLC patients, bsPD-L1 levels were almost unchanged during ICI treatment (Figure 1C and 1D), whereas anti-PD-L1, but not anti-PD-1, treatment increased sPD-L1 levels (Figure 1E). The sPD-L1 increase was associated with irAE development, but not with clinical outcomes (Figure 4). These results suggest that the sPD-L1 change can serve as an indicator to predict irAEs during anti-PD-L1 treatment.

Our results revealed the mechanism and source of sPD-L1 production during anti-PD-L1 treatment. Consistent with NSCLC patients, sPD-L1 levels were increased in mice treated with anti-PD-L1 mAb, but not with anti-PD-1 mAb. We observed trafficking of the anti-PD-L1 mAb to lysosomes in F4/80^+^ macrophages in various tissues (Figure 3B and 3C). Treatment with chloroquine inhibited the sPD-L1 increase induced by the anti-PD-L1 mAb, suggesting the involvement of lysosomal digestion in sPD-L1 production (Figure 3D). Furthermore, depletion of macrophages by clodronate-containing liposomes suppressed the anti-PD-L1-induced sPD-L1 increase (Figure 3E), suggesting that macrophages are the major source of sPD-L1 during anti-PD-L1 treatment. Our findings suggest that the mechanism of anti-tumor effect by anti-PD-L1 mAb is more likely to be degradation of PD-L1 rather than blockade of PD-1/PD-L1 signaling.

We also found that sPD-L1 production might be influenced by the tumor location. sPD-L1 was detected with much higher frequency in GC patients than in NSCLC patients (Figure 1A). We observed a stronger correlation between sPD-L1 and bsPD-L1 levels in GC patients than in NSCLC patients (Figure 1B). Because the gastrointestinal tract contains many digestive enzymes compared with lung tissues, it is likely that bsPD-L1 degradation by digestive enzymes contributes to sPD-L1 production.

bsPD-L1 levels were correlated with MMP13, MMP3, and IFN-γ levels (Figure 2A), while sPD-L1 levels were correlated with IL-1α, IL-1β, TNF-α, and IL-6 levels (Figure 2B). We speculate that MMP-mediated cleavage might be involved in bsPD-L1 production, whereas inflammation might be involved in sPD-L1 production. We have previously reported that glycosylation of bsPD-L1 is important for its binding to PD-1^17^. MMP-mediated PD-L1 cleavage may retain its conformational structure and glycosylated sites necessary for PD-1 binding. However, in inflamed tissues, PD-L1-expressing apoptotic/necrotic cells might be phagocytosed and digested in macrophage lysosomes^19^. Because lysosomes contain many hydrolytic enzymes, including proteases and glycosidases^20^, hydrolysis may degrade sugar chains, resulting in production of sPD-L1 without PD-1-binding capacity.

In this study, we demonstrated the association of sPD-L1 increase with irAE development, but not with clinical outcomes during anti-PD-L1 treatment (Figure 4). We have recently demonstrated that bsPD-L1 can function as an endogenous PD-1 blocker and serve as a biomarker for predicting the efficacy of ICIs (preprint in medRxiv/2023/300672). Unlike bsPD-L1, sPD-L1 without PD-1-binding ability does not have immunological functions to enhance T cell response. However, sPD-L1 production might reflect PD-L1 degradation in peripheral tissues, which causes a breakdown of peripheral tolerance leading to irAE development. Thus, the sPD-L1 change during anti-PD-L1 treatment may serves as an indicator to predict irAEs. Since sPD-L1 and bsPD-L1 have different clinical values, the combination of sPD-L1 and bsPD-L1 might represent a powerful non-invasive diagnostic tool to improve cancer immunotherapy by preventing side effects and stratifying patients.

## Data Availability

All data produced in the present work are contained in the manuscript.

## Acknowledgments

This work was partially supported by the Grant-in-Aid for Scientific Research from the Japan Society for the Promotion of Science (JP19K07783 and JP22K07262 to Y. I.) and a research grant from Sysmex Corporation (to Y. I.). We thank K. Nishimaki for technical and secretarial assistance.

## Authors’ contributions

Conceptualization: Y. I.; Methodology: Y. I., F. A., and Y. M.; Investigation: R. T., F. A., S. K., and Y. M.; Data analysis: Y. I., T. K., R. T., F. A., T. A., and M. H.; Resources: T. K., F. A., Y. K, H. Y., M. S., and A. G.; Funding acquisition: Y. I.; Supervision: Y. I. and A. G.; Writing – original draft: Y. I., T. K., and R. T.; Writing – review & editing: All authors.

## Disclosure of Potential Conflicts of Interest

Y. Iwai has patent applications for immunopotentiating compositions (WO/2009/0297518, 2011/0081341, 2014/0314714, 2015/0093380, 2015/0197572, 2016/0158356, 2016/0158355, 2017/0051060, and 2020/0062846) and an immune function evaluation method (WO/2019/049974). Y. Iwai has received research grants from the Japan Society for the Promotion of Science (JP19K07783 and JP22K07262) and Sysmex Corporation. A. Gemma reports consulting fees from MSD, Nippon Kayaku, and Daiichi-Sankyo Company outside the submitted work. M. Seike has received research grants from Taiho Pharmaceutical, Chugai Pharmaceutical, Eli Lilly, Nippon Kayaku, and Kyowa Hakko Kirin and honoraria from AstraZeneca, MSD, Chugai Pharmaceutical, Taiho Pharmaceutical, Eli Lilly, Ono Pharmaceutical, Bristol-Myers Squibb, Nippon Boehringer Ingelheim, Pfizer, Novartis, Takeda Pharmaceutical, Kyowa Hakko Kirin, Nippon Kayaku, Daiichi-Sankyo Company, Merck Biopharma, and Amgen outside the submitted work. The other authors declare no potential conflicts of interest.

## Notes

### Funding Statement

This work was supported by the Grant-in-Aid for Scientific Research from the Japan Society for the Promotion of Science (JP19K07783 and JP22K07262 to Y. I.) and a research grant from Sysmex Corporation (to Y. I.).

### Author Declarations

The study protocols (B-2019-005 and 28-09-646) were reviewed and approved by the Ethics Committee of Nippon Medical School.

