## Supplementary Fig.1 for "Lysosomal degradation of PD-L1 is associated with immune-related adverse events during anti-PD-L1 immunotherapy in NSCLC patients"

### Supplementary Figure 1

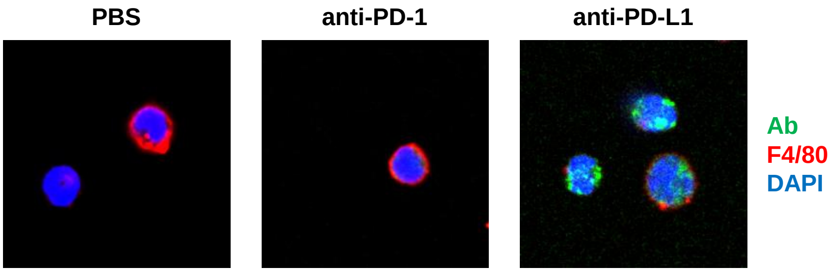


**Intracellular trafficking of anti-PD-L1 mAb in mouse bone marrow F4/80$^{+}$ macrophages.** Mice were intravenously injected with PBS, Alexa 488-labeled anti-PD-1 or anti-PD-L1 mAb (green). Bone marrow cells were isolated 24 h later and stained with anti-F4/80 mAb (red) and DAPI (blue).
