## Supplementary Table 1 for "Lysosomal degradation of PD-L1 is associated with immune-related adverse events during anti-PD-L1 immunotherapy in NSCLC patients"

**Supplementary Table 1** Patient characteristics

| Variable | | All patients (N=72) | anti-PD-1 (N=48) | anti-PD-L1 (N=24) | p value |
|---|---|---|---|---|---|
| ICI treatment | | | | | |
| | Nivolumab | 20 (27.8%) | 20 (41.6%) | | |
| | Pembrolizumab | 28 (38.9%) | 28 (58.3%) | | |
| | Atezolizumab | 16 (22.2%) | | 16 (66.7%) | |
| | Durvalumab | 8 (11.1%) | | 8 (33.3%) | |
| Age | Median (range) | 67 (28-88) | 70 (28-88) | 62 (43-77) | 0.0551 |
| Gender | | | | | 0.4227 |
| | Male | 56 (77.8%) | 36 (75.0%) | 20 (83.3%) | |
| | Female | 16 (22.2%) | 12 (25.0%) | 4 (16.7%) | |
| Smoking | | | | | 0.3542 |
| | Never | 11 (15.3%) | 6 (12.5%) | 5 (20.8%) | |
| | Current or Former | 61 (84.7%) | 42 (87.5%) | 19 (79.2%) | |
| EGFR Mutation | | | | | 0.7706 |
| | Wild | 60 (88.2%) | 42 (87.5%) | 18 (90.0%) | |
| | Mutant | 8 (11.8%) | 6 (12.5%) | 2 (10.0%) | |
| T | | | | | 0.2482 |
| | -1c | 18 (25.0%) | 14 (29.2%) | 4 (16.7%) | |
| | 2a- | 54 (75.0%) | 34 (70.8%) | 20 (83.3%) | |
| N | | | | | 0.2468 |
| | 0 | 11 (15.3%) | 9 (18.8%) | 2 (8.3%) | |
| | 1- | 61 (84.7%) | 39 (81.2%) | 22 (91.7%) | |
| M | | | | | 0.1528 |
| | 0 | 49 (68.1%) | 30 (62.5%) | 19 (79.2%) | |
| | 1 | 23 (31.9%) | 18 (37.5%) | 5 (20.8%) | |
| Stage | | | | | 0.0916 |
| | -II | 10 (13.9%) | 9 (18.8%) | 1 (4.2%) | |
| | III- | 62 (86.1%) | 39 (81.2%) | 23 (95.8%) | |
| Pathology | | | | | 0.2451 |
| | Squamous cell carcinoma | 32 (44.4%) | 24 (50.0%) | 8 (33.3%) | |
| | Adenocarcinoma | 32 (44.4%) | 18 (37.5%) | 14 (58.3%) | |
| | Non-small | 8 (11.2%) | 6 (12.5%) | 2 (8.4%) | |
| PD-L1 TPS status | | | | | 0.1500 |
| | <50 | 31 (52.5%) | 19 (46.3%) | 12 (66.7%) | |
| | 50≤ | 28 (47.5%) | 22 (53.7%) | 6 (33.3%) | |
